# Understanding inequalities in COVID-19 vaccination between migrants and non-migrants in Germany: The role of psychological factors of vaccine behaviour

**DOI:** 10.64898/2026.04.15.26350844

**Authors:** Susanne Bartig, Manuel Siegert, Claudia Hövener, Niels Michalski

## Abstract

**Background:** Understanding the underlying mechanisms for differences in vaccine uptake between migrants and non-migrants is crucial in order to design targeted interventions encouraging vaccination and to ensure vaccine-related equity. Therefore, this study examined to what extent migration-related disparities in COVID-19 vaccination were associated with psychological factors, based on the established 5C model of vaccine behaviour (Confidence, Complacency, Constraints, Calculation, Collective Responsibility).

**Methods:** Data were obtained from the German study “Corona Monitoring Nationwide – Wave 2” (RKI-SOEP-2 study), which was carried out between November 2021 and March 2022. The association between COVID-19 vaccination and migration status, while considering the psychological factors, was investigated using multivariable binary logistic regressions. A decomposition analysis (Karlson-Holm-Breen method) was conducted to examine the extent to which migration-related disparities in vaccine uptake were associated with the psychological factors of the 5C framework.

**Results:** Migrants were less likely to be vaccinated against COVID-19 compared to non-migrants, especially participants from the Middle East and North Africa (MENA) region. Our decomposition showed that almost two-thirds of the disparities in COVID-19 vaccine uptake between migrants and non-migrants were associated with the psychological factors (first-generation: 61.2%, second-generation: 64.2%). Confidence in safety of the vaccine was the most relevant factor in the 5C framework. Furthermore, the results highlighted the importance of a differentiated analysis regarding country of origin: While the 5C model accounted for only 19.4% of the difference between participants from the MENA region and non-migrants, the proportion for participants from Eastern Europe was 73.5%, suggesting that the underlying mechanisms for the lower uptake in the MENA group need further investigation.

**Conclusions:** Overall, migration-related disparities in COVID-19 vaccination were significantly associated with differences in psychological factors of vaccine behaviour. To increase vaccine acceptance within the heterogeneous group of migrants in general, tailored and proactive health communication interventions are needed.

## 1. Introduction

The outbreak of the novel coronavirus SARS-CoV-2 (severe acute respiratory syndrome coronavirus 2), which was declared a COVID-19 pandemic by the World Health Organization (WHO) in March 2020, has posed major challenges to societies worldwide. Vaccination against COVID-19 made a key contribution to protection against severe courses of the disease and helped contain the transmission of the virus throughout populations. In Germany, and in most countries worldwide, the COVID-19 vaccination was not mandatory but a free individual choice and free of charge. However, the vaccination campaign did not reach all population groups equally. Research shows that the likelihood of receiving the COVID-19 vaccines varied according to sociodemographic characteristics. For instance, groups with lower levels of education and income were less likely to be vaccinated [1]. Additionally, migrants and their (direct) descendants had lower vaccination rates compared to non-migrants [2]. Although several studies have addressed this issue, the reasons for the lower vaccination rates among migrants have not been conclusively determined. Migrants are often confronted with various barriers when accessing healthcare systems such as legal restrictions (especially asylum seekers), perceived discrimination, and language-related obstacles [3,4]. While language barriers may be less important for second-generation migrants, socio-economic disadvantages, social exclusion and lower health literacy can influence the utilization of healthcare services. These barriers can also affect access to vaccination services. A nationwide telephone-based survey in Germany indicated that beyond socioeconomic characteristics, perceived discrimination in the healthcare system and language barriers contributed to explaining the COVID-19 vaccination rate differences [5].

Beyond the above-described barriers influencing the accessibility of vaccination, psychological factors of vaccine behaviour could also explain disparities in vaccine uptake in general and also by migration status. The established 5C model provides a framework to explore individuals’ vaccine behaviour and is based on five ‘psychological antecedents of vaccination’ [6]: *Confidence* (e.g., trust in the safety and effectiveness of the vaccine, the health systems and policymakers), *Complacency* (perceived risk of a disease and whether vaccination is considered necessary), *Constraints* (structural and psychological barriers, including physical affordability, availability, ability to understand and geographical accessibility), *Calculation* (extent of individuals’ information-seeking and the evaluation of risks and benefits regarding vaccine uptake), *Collective responsibility* (willingness to protect others through vaccination) [6,7].

Research shows that migrants have more critical attitudes towards COVID-19 vaccination and higher vaccine hesitancy compared to non-migrants [8,9], like concerns about vaccine safety and side effects [9,10], or mistrust in government and health systems [11,12]. In Germany, only a few studies have investigated psychological factors of vaccine behaviour among migrants [13,14]. However, to our knowledge, no study has examined the extent to which psychological factors contribute to differences in COVID-19 vaccine uptake between migrants and non-migrants in Germany. Understanding the underlying mechanisms for the disparities is crucial for designing targeted interventions, such as communication programs that encourage vaccine uptake. These insights are important not only for managing future pandemics but also for combating vaccine hesitancy, improving vaccine acceptance, and thereby increasing vaccine uptake in general (e.g., for influenza). Hence, this study aimed to investigate the association between COVID-19 vaccination and migration status, as well as to explore whether this relationship was associated with the psychological factors of vaccine behaviour using the 5C model. As the focus of our study is on first-generation migrants, we introduced a further differentiation by considering the country of origin. We hypothesized that differences in the psychological factors of vaccine behaviour between migrants and non-migrants were associated with disparities in COVID-19 vaccine uptake and therefore addressed the following research questions:

1. Were the disparities in COVID-19 vaccine uptake between migrants and non-migrants associated with the psychological factors of vaccine behaviour?
2. To what extent were the disparities associated with the psychological factors of vaccine behaviour?
3. Which psychological factors were most strongly associated with the disparities in vaccine uptake between migrants and non-migrants?

## 2. Methods

### 2.1 Data

The analyses are based on data from the second wave of the German seroepidemiological study “Corona Monitoring Nationwide – Wave 2 (RKI-SOEP-2)”. This study was embedded in the Socio-Economic Panel (SOEP) – a nationwide dynamic cohort based on random samples of individuals living in private households across Germany [15]. All persons aged 14 years and older who participated in the SOEP survey wave in 2021 were invited to the RKI-SOEP-2 study (gross sample). In addition, the study includes the IAB-SOEP Migration Sample, which consists of immigrants arriving in Germany since 1995 [16], and the IAB-BAMF-SOEP Survey of Refugees, which targets refugees who entered Germany between 2013 and 2020 [17]. Data collection was carried out from 12 November 2021 to 6 March 2022. An invitation package was sent to each target person, containing both an individual invitation and study materials (e.g., questionnaire, blood self-sampling kit for capillary blood). Respondents could complete the self-administered questionnaire either in paper form or online. The questionnaire covered topics such as experienced SARS-CoV-2 infections, COVID-19 vaccination status and willingness to be vaccinated, general health as well as health behaviour. Based on information about the main languages used by the individuals invited to participate from previous waves of the SOEP survey, the study materials were sent out in seven different languages (German, Arabic, Farsi, English, Polish, Bulgarian, and Romanian). To increase participation in the study, a post-paid monetary incentive (10 euros for adults, 5 euros for adolescents) was announced, along with written notification of their laboratory results. All participants gave written informed consent to participate in this study which was approved by the Ethics Committee of the Berlin Chamber of Physicians (Eth-33/20) in compliance with the Declaration of Helsinki.

A total of 11,162 respondents aged 14 years and older participated in the RKI-SOEP-2 study. The response rate was 53.7% (Response Rate 6) according to the standards of the American Association for Public Opinion Research [18]. More details on the study design, data collection, and methods of the RKI-SOEP-2 study have been published elsewhere [19].

### 2.2 Measures

#### Outcome: COVID-19 vaccination (at least one dose)

Participants were asked about the number of vaccine doses they had received as well as the respective vaccination dates. In the following analyses, individuals were considered vaccinated if they had received at least one dose of a COVID-19 vaccine before the start of the survey (November 15, 2021).

#### Population groups: Migration status and country of origin

Migration status was categorized into “non-migrants”, “first-generation” (people who immigrated themselves), and “second-generation” migrants (at least one parent was not born in Germany). In this paper we used the term ‘migrants’ following the recommendations for analysing migration-related determinants in public health research [20]. To consider the heterogeneity of these populations, we used a subdivision regarding the country of origin (groups) for our analyses that was as differentiated as our data allowed. To this end, we distinguished among people with a personal migration experience (first-generation) who came from the “Middle East or Northern Africa (MENA)”, from extended “Eastern Europe” - including not only the classical Eastern European states but also countries of the Baltic states and Post-Soviet countries - and “other parts of the world”. The category “other parts of the world” consists of 6.4% Northern Europe, 37.3% Western Europe, 23.7% Southern Europe, 17.6% North and South America, 8.4% Asia, 6.1% Africa and 0.5% Australia and New Zealand.

#### Mediator: The 5C model

The psychological antecedents of vaccination (5Cs) were surveyed with one item for each concept: *confidence* (“I have complete trust in the safety of the COVID-19 vaccination”), c*omplacency* (“COVID-19 is not a major threat, thus vaccinations against COVID-19 are superfluous”), *calculation* (“When I think about getting vaccinated against COVID-19, I carefully weigh up the benefits and risks in order to make the best possible decision”), *collective responsibility* (“Vaccination is a community measure to prevent the spread of COVID-19”), and *constraints* (“It is too much hassle for me to get the COVID-19 vaccine”). The approval of each item was assessed on a 5-point Likert scale. We recoded the scale so that higher scores indicated stronger agreement, ranging from “does not apply at all” (1) to “applies completely” (5).

#### Confounding variables

We controlled for several sociodemographic characteristics that are potential confounders, as they have been shown in previous analyses to be associated with attitudes towards vaccination and vaccine uptake, while posing a low risk of acting as colliders. Specifically, this included *gender* (“female” and “male”), *age groups* (“18 to 30 years”, “40 to 59 years”, and “60 years or older”), *date of participation* in the survey, and *federal state* as well as *residential area* (“urban” vs. “rural”). According to the 2011 version of the International Standard Classification of Education (ISCED 2011 [21]), *education* was classified as “low” (ISCED 0-2), “medium” (3-4), and “high” (5-8) groups based on the educational and vocational qualifications of the study participants. *Income* was measured by equivalized monthly net household income, using the square-root method [22] and categorized into “low” (quintile 1), “medium” (quintile 2-4), and “high” (quintile 5). Besides the sociodemographic characteristics, *previous SARS-CoV-infection* before the field start of the survey (“yes” vs. “no”) and *selected chronic diseases* such as asthma, diabetes, cardiac disease (cardiac insufficiency, weak heart) as well as high blood pressure (“yes” vs. “no”) were considered as control variables.

### 2.3 Statistical analysis

To describe the sample characteristics, proportions and prevalences with 95% confidence intervals (95% CI) for COVID-19 vaccination were reported. Furthermore, descriptive analyses were conducted to explore differences between migrants and non-migrants across the psychological factors of COVID-19 vaccination. The results are presented as mean scores and their standard errors. The association between vaccine uptake and migration status, while considering the psychological factors, was estimated by calculating odds ratios (ORs) with 95% CI and p-values, using binary logistic regressions, with household-clustered standard errors. Adjustments were made for various sociodemographic and health-related characteristics (see “confounding variables”). Finally, a decomposition analysis was conducted to estimate the extent to which psychological factors accounted for migration-related disparities in COVID-19 vaccine uptake by using the Karlson-Holm-Breen (KHB) method [23]. Based on the Baron and Kenny approach [24], we explored whether the psychological factors could account for differences in vaccine uptake between population groups. While this approach is broadly in line with more general causal mediation frameworks it assumes no interaction between the exposure and mediator in their effects on the outcome (e.g., Valeri & VanderWeele 2013 [24]). Because our study compares distinct populations rather than variation in exposure within a single population, and migration cannot meaningfully be treated as an exposure within a causal (i.e. counterfactual) framework, we do not interpret the results as evidence of causal mediation. Instead, the analysis is intended to describe whether psychological factors may statistically account for the observed group differences. The rationale for using the Karlson-Holm-Breen (KHB) method concerns only the accommodation of non-linear models and otherwise does not differ from the standard mediation approach. The approach allowed us to determine to what extent the 5C framework accounted for the disparities in vaccine uptake between migrants and non-migrants by decomposing the associations into total, direct and indirect effects.

The analyses were calculated with weighting factors to compensate for systematic non-response and to adjust the sample to match the official German population statistics by age, sex, citizenship (German vs. non-German), federal state, household type and size, as well as owner-occupied housing [25]. In the analyses, respondents aged 18 years and older, as well as valid questionnaires were included (n=10,288). All analyses were conducted using Stata 17.0.

## 3. Results

### 3.1 Descriptive findings

A description of the study population by COVID-19 vaccination is presented in Table 1. Of the 10,288 participants aged 18 years and older, 93.2% reported having been vaccinated against COVID-19 at least once.

**Table 1.**
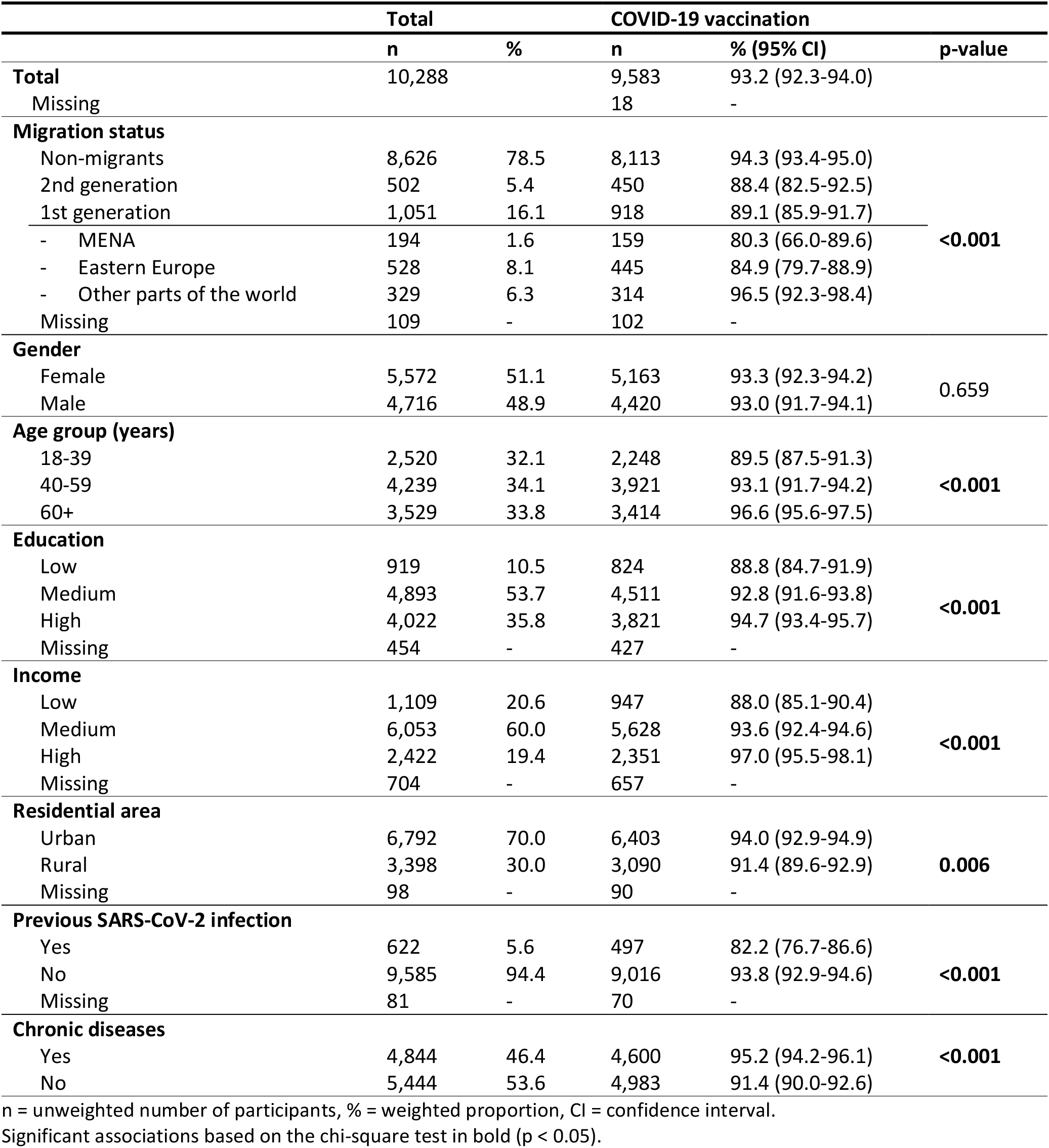
Description of the study population by COVID-19 vaccination.

The vaccination rate differed by migration status: The proportion of those who had been vaccinated at least once was lower among people who immigrated themselves (89.1%) and those of the second-generation (88.4%) compared to non-migrants (94.3%). Moreover, the results showed differences according to country of origin among people with a personal migration experience (first-generation). Whereas 80.3% of the participants from the MENA region and 84.9% from Eastern Europe were vaccinated against COVID-19, the vaccination rate for people who came from other parts of the world (96.5%) was at a similar level as non-migrants.

Table 2 indicates that the 5C drivers of vaccine behaviour differed by migration status. Non-migrants reported higher confidence in the safety of the COVID-19 vaccines and higher collective responsibility compared to first-generation migrants. In contrast, respondents who immigrated themselves showed a greater conviction that COVID-19 was not a major threat and vaccinations superfluous than non-migrants (complacency). A differentiated analysis by country of origin suggested that people from Eastern Europe had less confidence and collective responsibility regarding COVID-19 vaccination compared to participants from the other regions (Table S1).

**Table 2.**
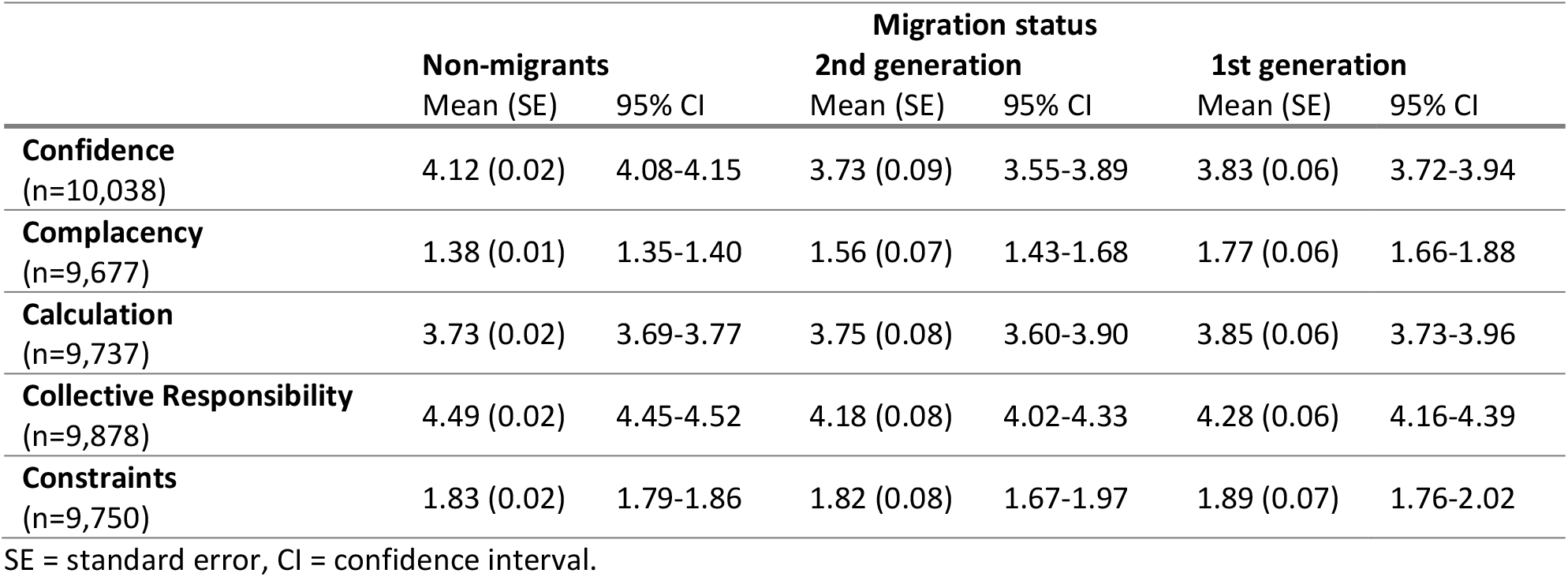
Descriptive statistics for study variables by migration status, mean (SE), and 95% CI, weighted.

### 3.2 Disparities in COVID-19 vaccination by migration status

In a next step, differences in COVID-19 vaccine uptake by migration status were analysed while considering the 5C framework. Results of the logistic regression models controlling for various sociodemographic and health-related characteristics are presented in Table 3. The base model (model 1) showed that people who immigrated themselves (OR=0.58; 95% CI 0.39-0.85; p=0.006) and whose parents had immigrated (OR=0.57; 95% CI 0.33-0.98; p=0.043) were less likely to be vaccinated against COVID-19 than non-migrants. The 5C drivers of vaccine behaviour were added separately in regression models 2 to 6. When confidence (model 2) and collective responsibility (model 5) were included, the effect of migration on vaccine uptake lost significance, suggesting that both psychological factors could explain the migration-related disparities in COVID-19 vaccination. For first-generation migrants, complacency also seemed to be important for the lower likelihood of being vaccinated compared to non-migrants.

**Table 3.** Logistic regression analyses predicting adjusted odds ratios of COVID-19 vaccination (n=9,526).

|  | (1) | (2) | (3) | (4) | (5) | (6) | (7) |
| --- | --- | --- | --- | --- | --- | --- | --- |
| Migration status<br>(Ref.: non-migrants) |  |  |  |  |  |  |  |
| 2nd generation | **0.57**<br>(0.043) | 0.63<br>(0.168) | **0.54**<br>(0.034) | **0.56**<br>(0.041) | 0.59<br>(0.125) | **0.58**<br>(0.044) | 0.75<br>(0.423) |
| 1st generation | **0.58**<br>(0.006) | 0.71<br>(0.184) | 1.04<br>(0.879) | **0.59**<br>(0.008) | 0.68<br>(0.145) | **0.58**<br>(0.008) | 0.68<br>(0.149) |
| Confidence | | **7.28**<br>($<0.001$) | | | | | **4.16**<br>($<0.001$) |
| Complacency | | | **0.33**<br>($<0.001$) | | | | 0.85<br>(0.138) |
| Calculation | | | | **0.58**<br>($<0.001$) | | | **0.64**<br>($<0.001$) |
| Collective Responsibility | | | | | **4.10**<br>($<0.001$) | | **1.86**<br>($<0.001$) |
| Constraints | | | | | | **1.56**<br>($<0.001$) | **1.41**<br>(0.001) |
| Pseudo $R^2$ | 0.11 | 0.53 | 0.28 | 0.14 | 0.44 | 0.13 | 0.58 |

OR = Odds ratios from logistic regressions with adjustments for age, gender, education, income, residential area, federal state, previous SARS-CoV-2 infection, chronic diseases, and date of participation. Bolded type indicates statistical significance ( $p < 0.05$ ); CI = confidence interval, Ref = reference group.

All psychological factors were associated with COVID-19 vaccination: Participants with higher confidence (model 2), collective responsibility (model 5), and constraints (model 6) were more likely to be vaccinated against COVID-19. Conversely, complacency (model 3), as well as calculation (model 4), were associated with a lower uptake of COVID-19 vaccine. In the final model (7), all 5C concepts were included and remained unchanged in terms of direction and significance, with the exception of complacency. Including confidence in model 2 increased the pseudo R^2^ substantially, while adding all psychological factors in the final model increased the pseudo R^2^ only marginally more.

Lastly, we explored whether the differences in COVID-19 vaccine uptake between migrants and non-migrants were associated with the psychological antecedents identified by the 5C framework (Table 4). The results of the KHB decomposition indicated that the 5Cs of vaccine behaviour were associated with disparities in COVID-19 vaccination: The psychological factors accounted for 61.2% of the difference in vaccine uptake for first-generation migrants and 64.2% for those of the second-generation, compared to non-migrants. Confidence in safety of the COVID-19 vaccination and collective responsibility showed the strongest associations within the 5C framework. Thus, confidence accounted for 40.1% respectively 41.4% of disparities in vaccine uptake between migrants and non-migrants.

**Table 4.** Results of the decomposition according to the Karlson-Holm-Breen (KHB) method by migration status (n=9,526).

| Migration status<br>(Ref.: non-migrants) | 2nd generation |  | 1st generation |  |
| --- | --- | --- | --- | --- |
| Effect | OR (95% CI) | p-value | OR (95% CI) | p-value |
| Total | 0.44 (0.22-0.91) | **0.026** | 0.37 (0.21-0.63) | **<0.001** |
| Direct | 0.75 (0.37-1.52) | 0.423 | 0.68 (0.40-1.15) | 0.149 |
| Indirect | 0.59 (0.38-0.94) | **0.025** | 0.54 (0.34-0.86) | **0.009** |
| Components | Diff. in % |  | Diff. in % |  |
| Confidence | 41.42 |  | 40.07 |  |
| Complacency | 1.77 |  | 5.83 |  |
| Calculation | 3.71 |  | 5.90 |  |
| Collective Responsibility | 14.54 |  | 12.48 |  |
| Constraints | 2.81 |  | -3.08 |  |
| Mediation in % | **64.24** |  | **61.20** |  |

Adjusted for age, gender, education, income, residential area, federal state, previous SARS-CoV-2 infection, chronic diseases, and date of participation. Bolded type indicates statistical significance ( $p < 0.05$ ); CI = confidence interval, Ref = reference group.

### 3.3 Disparities in COVID-19 vaccination by country of origin

Regarding country of origin, participants from the MENA region (OR=0.13; 95% CI 0.03-0.52; p=0.004) and Eastern Europe (OR=0.23; 95% CI 0.11-0.45; p<0.001) were less likely to be vaccinated compared to non-migrants (Table S2). Nonetheless, even after controlling for the 5C drivers of vaccine behaviour, significant differences between non-migrants and participants from the MENA region remained. In contrast, accounting for the psychological factors markedly reduced the disparities between second-generation migrants and non-migrants, as well as between participants from Eastern Europe and non-migrants, to the extent that the direct effects were no longer statistically significant. Our mediation analysis differentiated by country of origin highlighted significant variations in the role of the psychological factors (Table 5). While the 5C model accounted for 73.5% of the difference between participants from Eastern Europe and non-migrants, the proportion was only 19.4% for the MENA region. Although, the indirect effect for the MENA region was numerically notable, it was not statistically significant and certainly not strong enough to overcome the largest of disparities of any of the possible group comparisons. This suggested that drivers could be at work, which were different from those of the other (non-)migrant groups. For other parts of the world, the mediation analysis indicated no significant direct, indirect, or total effects (not shown).

**Table 5.** Results of the decomposition according to the Karlson-Holm-Breen (KHB) method by country of origin (n=9,526).

| Country of origin<br>(Ref.: non-migrants) | 2nd generation |  | MENA |  | Eastern Europe |  |
| --- | --- | --- | --- | --- | --- | --- |
| Effect | OR | p-value | OR | p-value | OR | p-value |
| Total | 0.43 (0.21-0.86) | **0.017** | 0.13 (0.03-0.52) | **0.004** | 0.23 (0.11-0.45) | **<0.001** |
| Direct | 0.73 (0.36-1.46) | 0.367 | 0.19 (0.05-0.77) | **0.020** | 0.68 (0.34-1.33) | 0.255 |
| Indirect | 0.59 (0.27-1.27) | 0.178 | 0.67 (0.31-1.46) | 0.315 | 0.34 (0.54-0.73) | **0.006** |
| Components | Diff. in % |  | Diff. in % |  | Diff. in % |  |
| Confidence | 40.09 |  | 1.87 |  | 50.12 |  |
| Complacency | 1.65 |  | 2.25 |  | 5.66 |  |
| Calculation | 3.76 |  | 9.09 |  | 4.06 |  |
| Collective Responsibility | 13.94 |  | 3.62 |  | 15.24 |  |
| Constraints | 2.86 |  | 2.54 |  | -1.58 |  |
| Mediation in % | 62.30 |  | 19.38 |  | **73.50** |  |

Adjusted for age, gender, education, income, residential area, federal state, previous SARS-CoV-2 infection, chronic diseases, and date of participation. Bolded type indicates statistical significance ( $p < 0.05$ ); CI = confidence interval, Ref = reference group.

## 4. Discussion

The COVID-19 pandemic reproduced and exacerbated health inequities between migrants and non-migrants insofar as migrants were at higher risk of SARS-CoV-2 infection and disproportionately affected by hospitalizations and deaths [26,27]. Regarding vaccine-related inequalities, this study (1) examined the association between migration and COVID-19 vaccine uptake as well as (2) explored the role of the psychological antecedents of vaccination in this relationship (5C model). In line with previous research, our results suggest disparities in COVID-19 vaccination between migrants and non-migrants [2]. People with a familial (second-generation) and personal history of migration (first-generation), especially from the MENA region, were less likely to be vaccinated against COVID-19 compared to non-migrants. In general, all psychological factors of vaccine behaviour were associated with COVID-19 vaccination, which is in line with other studies where the 5Cs related to disparities in COVID-19 vaccine uptake [28,29]. Confidence in the safety of COVID-19 vaccination and collective responsibility increased the likelihood of being vaccinated. Moreover, perceived constraints were positively associated with vaccination. This unexpected result may be attributable to reverse causation or reporting bias, as only people who intended to, or had already been vaccinated, could perceive and report such access barriers. In contrast, the conviction that vaccinations are superfluous (complacency) and a careful cost-risk analysis (calculation) were associated with a lower chance of being vaccinated.

The underlying mechanisms for the differences in COVID-19 vaccination between migrants and non-migrants remain insufficiently researched and have focused on structural barriers to access, such as language requirements or experiences of discrimination. Hence, this study explored to what extent psychological factors of vaccine behaviour, utilizing the 5C model as a framework, were associated with disparities in COVID-19 vaccine uptake between migrants and non-migrants, closing a major research gap. The results of the decomposition analysis suggest that the psychological factors account for a substantial proportion of the differences in COVID-19 vaccine uptake. Thus, almost two-thirds of the effect of migration on COVID-19 vaccination was associated with the psychological factors. Overall, confidence in the safety of the vaccination emerged as the strongest psychological factor associated with disparities in vaccine uptake.

The psychological factor ‘confidence’ is influenced by knowledge about COVID-19 [8], suggesting one explanation for differences in vaccine uptake by migration status. A German telephone survey showed that uncertainties and false knowledge about the vaccination were more common among migrants than among non-migrants [5]. Furthermore, own calculations based on the RKI-SOEP-2 study showed that the perceived informedness about COVID-19 vaccination was lower among first-generation migrants compared to non-migrants. At the beginning of the pandemic, adequate official information was not available in the languages spoken by the migrants [30]. The lack of accessible, trustworthy, and translated information can lead to language barriers, which in turn promote the use of alternative sources of information and thereby increase susceptibility to misinformation, often spread through social media [26]. A rapid review has suggested that misinformation about COVID-19 vaccines circulating on social media contributed to vaccine hesitancy [31]. A lack of confidence resulting from social media use is therefore another possible explanation for the differences in vaccination. Own calculations based on the RKI-SOEP-2 study also indicated that first- and second-generation migrants used social media more frequently as information source than non-migrants (Table S3). Among first-generation migrants, the social media use varied according to the length of stay. Participants who lived in Germany for up to 10 years used social media most frequently. Holz et al. (2022) showed an association between ‘years since migration’ and German media consumption, which mediated trust and thus the intention to get vaccinated [13]. The duration of residence was found to be an important factor influencing COVID-19 vaccine uptake in different studies [32,33]. It is likely that increasing duration of residence is accompanied with a better understanding of the host country healthcare system and language skills, which strengthen health literacy. Research has suggested an effect of health literacy on vaccine hesitancy [34] and has indicated a mediating role of the psychological antecedents in this association [35].

Institutional trust is another important factor that can reduce the negative effects of misinformation on vaccine hesitancy and uptake [36]. ‘Trust in government’ turned out to be a bridging factor linked with vaccine confidence attitudes, which were directly associated with vaccine uptake [37]. In addition to (mis-)trust in government, vaccine hesitancy can also relate to mistrust in the healthcare system [38], arising from structural inequities and racial discrimination [4]. Thus, previous experiences of discrimination in health services can have a negative impact on trust and consequently reduce the willingness to get vaccinated. Studies have indicated that experiences of racial as well as ethnic discrimination can increase COVID-19 vaccine hesitancy [39,40] and that mistrust in health systems can mediate this effect [41]. Experiences of racial discrimination and institutional mistrust can therefore represent further explanatory factors for the identified major importance of confidence in explaining the differences in vaccination. In addition, experiences of discrimination can influence the sense of belonging to the society in Germany, which is lower among migrants than among non-migrants [42], and therefore decrease collective responsibility regarding COVID-19 vaccination. To summarize, differences in vaccine uptake, related to lower levels of confidence and collective responsibility among migrants, were rooted in structural inequities, such as experiences of racial discrimination and misinformation about vaccines, which can also influence complacency.

First-generation migrants and their descendants are a heterogeneous population. Our results highlight the relevance of a differentiated analysis regarding country of origin. While psychological factors played a significant role in the vaccination behaviour of respondents from Eastern Europe, there is a need to identify the underlying mechanisms for the lower vaccination rate of respondents from the MENA region. Previous research showed that people from Eastern Europe were more likely vaccine hesitant for infectious diseases in general, such as measles or influenza [3]. Furthermore, a systematic review found that the COVID-19 vaccine acceptance rates was lowest among people from Eastern Europe, which may be related to lower confidence in the safety and effectiveness of the vaccines [43] and lower trust in the health system [44]. Regarding the respondents from the MENA region, of whom the vast majority have a refugee history and a shorter length of residence than respondents from Eastern Europe, it can be assumed that structural inequalities in accessing health services are more likely to explain differences in vaccine uptake than the psychological factors. Many general practitioners do not know that the Asylum Seekers’ Benefits Act includes vaccinations in Germany [14]. Besides structural barriers such as language-related barriers, refugees face concerns about their residency status. In a qualitative study in the United Kingdom, fear of being charged for vaccination, distrust in medical organizations, and worries of being asked about one’s legal status were identified as potential barriers of vaccination [45].

### 4.1 Strengths and limitations

To our knowledge, this was the first study to investigate the extent to which the psychological antecedents of vaccination (5Cs) accounted for disparities in COVID-19 vaccine uptake between migrants and non-migrants in Germany. A major strength of the RKI-SOEP-2 study was the design and sampling which included special samples of migrants and refugees to reflect the diversity in Germany. Furthermore, the use of multilingual study and survey materials ensured that the non-German-speaking population was reached in the study. Another strength of the study was its integration in the long-running dynamic cohort SOEP, which enabled both differentiated analyses according to a variety of sociodemographic as well as health-related characteristics and to compensate for possible bias due to systematic non-participation through weighting for a higher generalizability to the general population.

However, some limitations should be noted. Given the cross-sectional design of the RKI-SOEP-2 study, temporal ordering cannot be established and causal interpretations are therefore not warranted. The findings of the mediation analyses should be interpreted carefully, as cross-sectional approaches may produce biased estimates compared to longitudinal designs [46]. In particular, since the psychological antecedents of vaccination (5Cs) are measured after the COVID-19 vaccination, reverse causality cannot be ruled out. Vaccination status may also shape psychological attitudes toward vaccination. Unfortunately, longitudinal data were not available to establish causal relationships for the measures and to better understand the temporal dynamics. However, mediation analysis still helps to shed light on potential mechanisms which may underlie the observed associations.

In addition, the response rates were lower among migrants. One possible explanation could be that the regular SOEP waves are usually conducted as face-to-face interviews. Due to the pandemic, the study did not involve personal contact but instead relied on a written format or used the ‘computer-assisted web interviewing’ (CAWI) survey technique. Another limitation was selection effects caused by differences in willingness to participate, which could not be compensated by weighting and may result in sample bias. Vaccinated people or those with more positive attitudes towards vaccination were more likely to participate in this survey, which may have led to an overestimation of the vaccination rate [47]. The difference between the proportion of vaccinated people (at least one dose) in our survey sample (93.2%) and the official vaccination rate, which was reported at around 75% at the end of 2021, supports this assumption [48]. Lastly, the 5Cs were measured using single items, which is a common approach in vaccine-related research. While the items of the single-item version of the 5C short scale are chosen to best represent each construct [6], multi-item measures would have further strengthened the psychometric robustness of our results. Overall, despite these limitations, our findings provide important insights regarding the underlying mechanisms for migration-related disparities in COVID-19 vaccination in Germany in order to design targeted strategies to overcome vaccine hesitancy.

## 5. Conclusions

Considering the socially unequal distribution of SARS-CoV-2 infection risk and the severe courses of COVID-19, the vaccine-related inequities indicated in the present study highlight the need for health-policy measures. In order to design targeted interventions to increase vaccine acceptance, it is necessary to identify the factors associated with vaccine-related inequities. Based on the identification of disparities in COVID-19 vaccination between migrants and non-migrants, we analysed the underlying mechanisms that generate inequalities in COVID-19 vaccine uptake focusing on psychological factors. As confidence emerged as the most relevant psychological factor, proactive health communication strategies to improve vaccine-related knowledge are needed. To ensure equitable access to vaccination for migrants, low-threshold, lifeworld-oriented, and targeted vaccination services – adapted to people’s living conditions and accompanied by proactive offers (outreach) – are required. This should include the involvement of (multilingual) mediators or key people from the communities to both disseminate information about the services and increase trust and acceptance of vaccinations. Furthermore, vaccine-related information must be accessible in the most commonly spoken languages. Against the background of the lower immunity coverage, for example, measles or influenza among migrants [49], and the high relevance of the 5Cs in explaining vaccine hesitancy [50], more in-depth research is needed with a health equity focus to explore the effect of structural inequities on the psychological factors of vaccine behaviour. In addition, further research is needed to evaluate which measures are effective in increasing vaccination acceptance within the heterogeneous group of migrants and their descendants.

## Supplementary Information

**Table S1** Descriptive statistics for study variables by country of origin, mean (SD) and 95% CI, weighted.

**Table S2** Logistic regression analyses predicting adjusted odds ratios of COVID-19 vaccination by country of origin (n=9,526).

**Table S3** Proportion of respondents who often used social media to get information about the coronavirus and the pandemic by migration status and duration of stay (n=9,903).

## Acknowledgements

We would like to thank our colleagues from the RKI-SOEP Study Group, including those at the Robert Koch Institute (RKI), the Socio-Economic Panel (SOEP) at the German Institute for Economic Research (DIW Berlin), the Institute for Employment Research (IAB) and the Research Centre of the Federal Office for Migration and Refugees (BAMF-FZ) with whom we jointly conducted the “Corona Monitoring Nationwide” (RKI-SOEP-2) stud. Special thanks go to the staff of DIW for carrying out the weighting of the data. We also thank the employees of the ‘infas – Institute for Applied Social Sciences’ who contributed to the planning and implementation of fieldwork and data collection. Finally, we sincerely thank all study participants for their willingness to participate.

## Author Contributions

Conceptualization: Susanne Bartig.

Methodology: Susanne Bartig, Niels Michalski.

Data analysis: Susanne Bartig.

Writing—original draft: Susanne Bartig.

Writing – review & editing: Susanne Bartig, Manuel Siegert, Claudia Hövener, Niels Michalski.

Supervision: Manuel Siegert, Claudia Hövener, Niels Michalski.

All authors have read and agreed to the published version of the manuscript.

## Data availability

For empirical analysis we used data from the “Corona-Monitoring Nationwide” survey in Germany from winter 2021/22. This survey is part of the German Socio-Economic Panel (SOEP). Data access is granted on request for scientific purpose on a public repository via a low-level data use request, described in the SOEP Research Data Centre (https://www.diw.de/en/diw_01.c.601584.en/data_access.html).

## Ethics approval and consent to participate

The study was approved by the Ethics Committee of the Berlin Chamber of Physicians (Eth-33/20) in compliance with the Declaration of Helsinki. All participants gave written informed consent.

## References

1. Bartig S, Beese F, Wachtler B, Grabka MM, Mercuri E, Schmid L, et al. Socioeconomic Differences in SARS-CoV-2 Infection and Vaccination in Germany: A Seroepidemiological Study After One Year of COVID-19 Vaccination Campaign. Int J Public Health. 2023;68:1606152.

2. Gualdi-Russo E, Zaccagni L. COVID-19 Vaccination and Predictive Factors in Immigrants to Europe: A Systematic Review and Meta-Analysis. Vaccines. 2024;12(4):350.

3. Crawshaw AF, Farah Y, Deal A, Rustage K, Hayward SE, Carter J, et al. Defining the determinants of vaccine uptake and undervaccination in migrant populations in Europe to improve routine and COVID-19 vaccine uptake: a systematic review. Lancet Infect Dis. 2022;22(9):e254–66.

4. Njoku A, Joseph M, Felix R. Changing the Narrative: Structural Barriers and Racial and Ethnic Inequities in COVID-19 Vaccination. Int J Environ Res Public Health. 2021;18(18):9904.

5. Wulkotte E, Schmid-Küpke N, Bozorgmehr K, Razum O, Wichmann O, Neufeind J. Barriers and drivers to COVID-19 vaccination among the migrant and non-migrant population in Germany, 2021. Eur J Public Health. 2024;34(3):530–6.

6. Betsch C, Schmid P, Heinemeier D, Korn L, Holtmann C, Böhm R. Beyond confidence: Development of a measure assessing the 5C psychological antecedents of vaccination. PLoS ONE. 2018;13(12):e0208601.

7. MacDonald NE. Vaccine hesitancy: Definition, scope and determinants. Vaccine. 2015;33(34): 4161–4.

8. Paul E, Steptoe A, Fancourt D. Attitudes towards vaccines and intention to vaccinate against COVID-19: Implications for public health communications. Lancet Reg Health Eur. 2021;1:100012.

9. Hajissa K, Mutiat HA, Kaabi NA, Alissa M, Garout M, Alenezy AA, et al. COVID-19 Vaccine Acceptance and Hesitancy among Migrants, Refugees, and Foreign Workers: A Systematic Review and Meta-Analysis. Vaccines. 2023;11(6):1070.

10. Lin S. COVID-19 Pandemic and Im/migrants’ Elevated Health Concerns in Canada: Vaccine Hesitancy, Anticipated Stigma, and Risk Perception of Accessing Care. J Immigr Minor Health. 2022;24(4):896–908.

11. Abba-Aji M, Stuckler D, Galea S, McKee M. Ethnic/racial minorities’ and migrants’ access to COVID-19 vaccines: A systematic review of barriers and facilitators. J Migr Health. 2022;5:100086.

12. Yazdani Y, Pai P, Sayfi S, Mohammadi A, Perdes S, Spitzer D, et al. Predictors of COVID-19 vaccine acceptability among refugees and other migrant populations: A systematic scoping review. PloS ONE. 2024;19(7):e0292143.

13. Holz M, Mayerl J, Andersen H, Maskow B. How Does Migration Background Affect COVID-19 Vaccination Intentions? A Complex Relationship Between General Attitudes, Religiosity, Acculturation and Fears of Infection. Front Public Health. 2022;10:854146.

14. Führer A, Pacolli L, Yilmaz-Aslan Y, Brzoska P. COVID-19 Vaccine Acceptance and Its Determinants among Migrants in Germany - Results of a Cross-Sectional Study. Vaccines. 2022;10(8):1350.

15. Goebel J, Grabka MM, Liebig S, Kroh M, Richter D, Schröder C, et al. The German Socio-Economic Panel (SOEP). J Econ Stat. 2019;239(2):345–60.

16. Brücker H, Kroh M, Bartsch S, Goebel J, Kühne S, Liebau E, et al. The new IAB-SOEP Migration Sample: an introduction into the methodology and the contents. In: SOEP Survey Papers 216: Series C. 2014. https://www.diw.de/documents/publikationen/73/diw_01.c.570700.de/diw_ssp0216.pdf. Accessed 13 March 2025.

17. Kühne S, Jacobsen J, Kroh M. Sampling in Times of High Immigration: The Survey Process of the IAB-BAMF-SOEP Survey of Refugees. Survey Methods: Insights from the Field. 2019. Available from: https://surveyinsights.org/?p=11416. Accessed 13 March 2025.

18. American Association for Public Opinion Research. Standard definitions: final dispositions of case codes and outcome rates for surveys. 9th ed. AAPOR; 2016.

19. Bartig S, Brücker H, Butschalowsky H, Danne C, Gößwald A, Goßner L, et al. Corona Monitoring Nationwide (RKI-SOEP-2): Seroepidemiological Study on the Spread of SARS-CoV-2 Across Germany. J Econ Stat. 2023;243(3-4):431–49.

20. Kajikhina K, Koschollek C, Sarma N, Bug M, Wengler A, Bozorgmehr K, et al. Recommendations for collecting and analysing migration-related determinants in public health research. J Health Monit. 2023;8(1):52–72.

21. UNESCO Institute for Statistics. International Standard Classification of Education: ISCED 2011. Montreal: UNESCO Institute for Statistics; 2012.

22. Atkinson A, Rainwater L, Smeeding T. Income distribution in OECD countries: evidence from the Luxembourg Income Study (LIS). Paris: OECD Publishing; 1995.

23. Kohler U, Karlson KB, Holm A. Comparing Coefficients of Nested Nonlinear Probability Models. The Stata Journal. 2011;10:420–38.

24. Valeri L, VanderWeele TJ. Mediation analysis allowing for exposure-mediator interactions and causal interpretation: theoretical assumptions and implementation with SAS and SPSS macros. Psychol Methods 2013;18(2):137–50.

25. Danne C, Priem M, Steinhauer H. SOEP-Core – 2021: Sampling, Nonresponse, and Weighting in Wave 2 of Living in Germany – Nationwide Corona-Monitoring (RKI-SOEP 2). In: SOEP Survey Papers: Series C. 2022. http://hdl.handle.net/10419/261439. Accessed 13 March 2025.

26. European Centre for Disease Prevention and Control. Reducing COVID-19 transmission and strengthening vaccine uptake among migrant populations in the EU/EEA. Available from: https://migrant-integration.ec.europa.eu/sites/default/files/2021-06/covid-19-reducing-transmission-and-strengthening-vaccine-uptake-in-migrants.pdf. Accessed 13 March 2025.

27. Hintermeier M, Gottlieb N, Rohleder S, Oppenberg J, Baroudi M, Pernitez-Agan S, et al. COVID-19 among migrants, refugees, and internally displaced persons: systematic review, meta-analysis and qualitative synthesis of the global empirical literature. eClinicalMedicine. 2024;74:102698.

28. Hossain MB, Alam MZ, Islam MS, Sultan S, Faysal MM, Rima S, et al. Health Belief Model, Theory of Planned Behavior, or Psychological Antecedents: What Predicts COVID-19 Vaccine Hesitancy Better Among the Bangladeshi Adults? Front Public Health. 2021;9:711066.

29. Rancher C, Moreland AD, Smith DW, Cornelison V, Schmidt MG, Boyle J, et al. Using the 5C model to understand COVID-19 vaccine hesitancy across a National and South Carolina sample. J Psychiatr Res. 2023;160:180–186.

30. Nezafat Maldonado BM, Collins J, Blundell HJ, Singh L. Engaging the vulnerable: A rapid review of public health communication aimed at migrants during the COVID-19 pandemic in Europe. J Migr Health. 2020;1-2:100004.

31. Skafle I, Nordahl-Hansen A, Quintana DS, Wynn R, Gabarron E. Misinformation About COVID-19 Vaccines on Social Media: Rapid Review. J Med Internet Res. 2022;24(8):e37367.

32. Bastola K, Nohynek H, Lilja E, Castaneda A E, Austero S, Kuusio H, et al. Incidence of SARS-CoV-2 Infection and Factors Associated With Complete COVID-19 Vaccine Uptake Among Migrant Origin Persons in Finland. Int J Public Health. 2023;68:1605547.

33. Diaz E, Dimka J, Mamelund SE. Disparities in the offer of COVID-19 vaccination to migrants and non-migrants in Norway: a cross sectional survey study. BMC Public Health. 2022;22(1):1288.

34. Montagni I, Ouazzani-Touhami K, Mebarki A, Texier N, Schück S, Tzourio C. Acceptance of a Covid-19 vaccine is associated with ability to detect fake news and health literacy. J Public Health. 2021;43(4):695–702.

35. Lu Y, Wang Q, Zhu S, Xu S, Kadirhaz M, Zhang Y, et al. Lessons learned from COVID-19 vaccination implementation: How psychological antecedents of vaccinations mediate the relationship between vaccine literacy and vaccine hesitancy. Soc Sci Med. 2023;336:116270.

36. Chen X, Lee W, Lin F. Infodemic, Institutional Trust, and COVID-19 Vaccine Hesitancy: A Cross-National Survey. Int J Environ Res Public Health. 2022;19(13):8033.

37. Yuan J, Xu Y, Wong IOL, Lam WWT, Ni MY, Cowling BJ, et al. Dynamic predictors of COVID-19 vaccination uptake and their interconnections over two years in Hong Kong. Nat Commun. 2024;15(1):290.

38. Charura D, Hill AP, Etherson ME. COVID-19 Vaccine Hesitancy, Medical Mistrust, and Mattering in Ethnically Diverse Communities. J Racial and Ethn Health Disparities. 2023;10(3):1518–25.

39. Savoia E, Piltch-Loeb R, Goldberg B, Miller-Idriss C, Hughes B, Montrond A, et al. Predictors of COVID-19 Vaccine Hesitancy: Socio-Demographics, Co-Morbidity, and Past Experience of Racial Discrimination. Vaccines. 2021;9(7):767.

40. Willis DE, Montgomery BEE, Selig JP, Andersen JA, Shah SK, Li J, et al. COVID-19 vaccine hesitancy and racial discrimination among US adults. Prev Med Rep. 2023;31:102074.

41. Paul E, Fancourt D, Razai M. Racial discrimination, low trust in the health system and COVID-19 vaccine uptake: a longitudinal observational study of 633 UK adults from ethnic minority groups. J R Soc Med. 2022;115(11):439–47.

42. Federal Statistical Office of Germany. Dashboard Integration. Available from: https://www.dashboard-integration.de/integration/zugehoerigkeitsgefuehl. Accessed 13 March 2025.

43. Sallam M. COVID-19 Vaccine Hesitancy Worldwide: A Concise Systematic Review of Vaccine Acceptance Rates. Vaccines. 2021;9(2):160.

44. Madar AA, Benavente P, Czapka E, Herrero-Arias R, Haj-Younes J, Hasha W, et al. COVID-19: information access, trust and adherence to health advice among migrants in Norway. Arch Public Health. 2022;80(1):15.

45. Deal A, Hayward SE, Huda M, Knights F, Crawshaw AF, Carter J, et al. Strategies and action points to ensure equitable uptake of COVID-19 vaccinations: A national qualitative interview study to explore the views of undocumented migrants, asylum seekers, and refugees. J Migr Health. 2021;4:100050.

46. Maxwell SE, Cole DA, Mitchell MA. Bias in Cross-Sectional Analyses of Longitudinal Mediation: Partial and Complete Mediation Under an Autoregressive Model. Multivariate Behav Res. 2011;46(5):816–41.

47. Mercuri E, Schmid L, Poethko-Müller C, Schlaud M, Kußmaul S, Ordonez-Cruickshank A, et al. Nationwide population-based infection- and vaccine-induced SARS-CoV-2 antibody seroprevalence in Germany in autumn/winter 2021/2022. Euro Surveill. 2025;30(1):2400037.

48. Steffen A, Rieck T, Fischer C, Siedler A. COVID-19 vaccine uptake - A special analysis with data up to December 2021. [Inanspruchnahme der COVID-19-Impfung – Eine Sonderauswertung mit Daten bis Dezember 2021]. Epid Bull. 2022;27:3–21.

49. Cherri Z, Lau K, Nellums LB, Himmels J, Deal A, McGuire E, et al. The immune status of migrant populations in Europe and implications for vaccine-preventable disease control: a systematic review and meta-analysis. J Travel Med. 2024;31(6):aae033

50. Eitze S, Felgendreff L, Horstkötter N, Seefeld L, Betsch C. Exploring pre-pandemic patterns of vaccine decision-making with the 5C model: results from representative surveys in 2016 and 2018. BMC Public Health. 2024;24(1):1205.

