## Supplementary Information for "Understanding inequalities in COVID-19 vaccination between migrants and non-migrants in Germany: The role of psychological factors of vaccine behaviour"

**Table S1** Descriptive statistics for study variables by country of origin, mean (SD) and 95% CI, weighted.

|  | **Country of origin (1^st^ generation)** | | | | | |
| --- | --- | --- | --- | --- | --- | --- |
|  | **MENA** | | **Eastern Europe** | | **Other parts of the world** | |
|  | Mean (SE) | 95% CI | Mean (SE) | 95% CI | Mean (SE) | 95% CI |
| **Confidence**  (n=10,038) | 3.96 (0.12) | 3.72-4.20 | 3.55 (0.08) | 3.39-3.71 | 4.15 (0.08) | 3.99-4.30 |
| **Complacency**  (n=9,677) | 1.92 (0.13) | 1.66-2.18 | 1.96 (0.08) | 1.80-2.13 | 1.49 (0.08) | 1.34-1.65 |
| **Calculation**  (n=9,737) | 3.98 (0.17) | 3.64-4.32 | 3.88 (0.08) | 3.72-4.03 | 3.79 (0.10) | 3.59-3.98 |
| **Collective Responsibility**  (n=9,878) | 4.14 (0.15) | 3.86-4.43 | 4.09 (0.09) | 3.92-4.27 | 4.53 (0.07) | 4.39-4.67 |
| **Constraints**  (n=9,750) | 1.64 (0.14) | 1.37-1.91 | 1.87 (0.09) | 1.69-2.05 | 1.97 (0.11) | 1.75-2.18 |

SE = standard error, CI = confidence interval

**Table S2** Logistic regression analyses predicting adjusted odds ratios of COVID-19 vaccination by country of origin (n=9,526).

|  | **Model 1**  **(reduced)** | | **Model 2**  **(full)** | |
| --- | --- | --- | --- | --- |
|  | OR (95% CI) | p-value | OR (95% CI) | p-value |
| Migration (Ref.: non-migrants) |  |  |  |  |
| 2^nd^ generation | 0.43 (0.21-0.86) | **0.017** | 0.73 (0.36-1.46) | 0.367 |
| Country of origin (1^st^ generation) |  |  |  |  |
| - MENA | 0.13 (0.03-0.52) | **0.004** | 0.19 (0.05-0.77) | **0.020** |
| - Eastern Europe | 0.23 (0.11-0.45) | **<0.001** | 0.68 (0.34-1.33) | 0.255 |
| Other parts of the world | 1.14 (0.40-3.28) | 0.810 | 1.23 (0.43-3.53) | 0.700 |
| Confidence |  |  | 4.24 (3.39-5.31) | **<0.001** |
| Complacency |  |  | 0.86 (0.69-1.06) | 0.159 |
| Calculation |  |  | 0.64 (0.53-0.77) | **<0.001** |
| Collective Responsibility |  |  | 1.86 (1.54-2.25) | **<0.001** |
| Constraints |  |  | 1.40 (1.15-1.72) | **<0.001** |
| Pseudo R^2^ | 0.11 |  | 0.59 |  |

OR = Odds ratios from logistic regressions with adjustments for age, gender, education, income, residential area, federal state, previous SARS-CoV-2 infection, chronic diseases, and date of participation. Bolded type indicates statistical significance (p < 0.05); reduced indicates that OR’s have been rescaled according to the full model using the KHB-method; CI = confidence interval, Ref = reference group.

**S3 Table** Proportion of respondents who often used social media to get information about the coronavirus and the pandemic by migration status and duration of stay (n=9,903).

|  | **n** | **%**  **(95% CI)** | **p-value** |
| --- | --- | --- | --- |
| **Migration status** |  |  | **<0.001** |
| Non-migrants | 1,034/8,468 | 12.7  (11.7-13.8) |  |
| 2nd generation | 96/493 | 22.5  (17.5-28.5) |  |
| 1st generation | 242/942 | 20.2  (16.8-24.1) |  |
| **Duration of stay** |  |  |  |
| Up to 10 years | 111/274 | 30.7  (23.1-39.6) | **0.006** |
| 11 to 20 years | 37/195 | 17.4  (11.3-25.7) |  |
| 21 years or more | 94/473 | 17.5  (13.1-22.9) |  |

n = unweighted number of participants, % = weighted proportion, CI = confidence interval. Significant associations based on the chi-square test in bold (p < 0.05).
